# Dynamics of RT-qPCR SARS-CoV-2 Detection Rates Prior to and After Symptom Onset

**DOI:** 10.1101/2020.07.09.20149245

**Authors:** Scott Sherrill-Mix

**Affiliations:** University of Pennsylvania

## Abstract

Effective RT-qPCR testing for SARS-CoV-2 is essential for treatment, surveillance and control of the COVID-19 pandemic. A recent meta-analysis ^1^ suggested that testing prior to the onset of symptoms is likely to miss the majority of infected individuals. These findings cast severe doubts on the effectiveness of mass screening efforts intended to detect SARS-CoV-2 prior to the onset of symptoms and decrease community transmissions from pre-/asymptomatic individuals^2–4^. However, alternative analyses and additional data described herein refine these estimates and suggest that many SARS-CoV-2 infections could potentially be detected prior to symptom onset.

## Main

In a recently published study, Kucirka et al. ^1^ collected data from seven studies of RT-qPCR testing of patients infected with SARS-CoV-2^5–11^ and concluded that detection of SARS-COV-2 was very difficult prior to symptom onset and effectively impossible earlier than three days prior to symptom onset. However the polynomial logistic regression model used for this analysis has the potential to underestimate uncertainty in time periods with sparse data and unfortunately only three tests from a single patient were available prior to the onset of symptoms. In order to more adequately account for this uncertainty, we developed a Bayesian autoregressive moving average state space model (B-ARMA-SSM) and used Markov chain Monte Carlo sampling to estimate posterior probabilities for parameters of interest from the data collected by Kucirka et al. ^1^

For the day of and days following symptom onset, the detection rates estimated by the B-ARMA-SSM and the polynomial model of Kucirka et al. ^1^ closely resembled each other (Figure 1). Both models estimated a peak in detection rates 3–4 days after symptom onset followed by a progressive decline in the probability of a positive detection. However in the days prior to symptom onset, the two models differed markedly in their estimates of detection rates and in their confidence in these estimates. In spite of the data only containing three tests in this time period, the polynomial model estimated very low detection rates, ostensibly precisely and with little potential for error. In contrast, the B-ARMA-SSM estimated higher probabilities of presymptomatic detection while displaying a much larger amount of uncertainty in its estimates (Figure 1). For example, at 4 days prior to symptom onset the B-ARMA-SSM estimates a detection rate of 35% (95% credible interval: 6–76%) while the polynomial model estimates a detection rate of 0% (95% CrI: 0–0%). Intuitively, the larger confidence intervals seem more appropriate given the limited testing available but without further data it is unclear which of these estimates is more likely to reflect reality.

**Figure 1:**
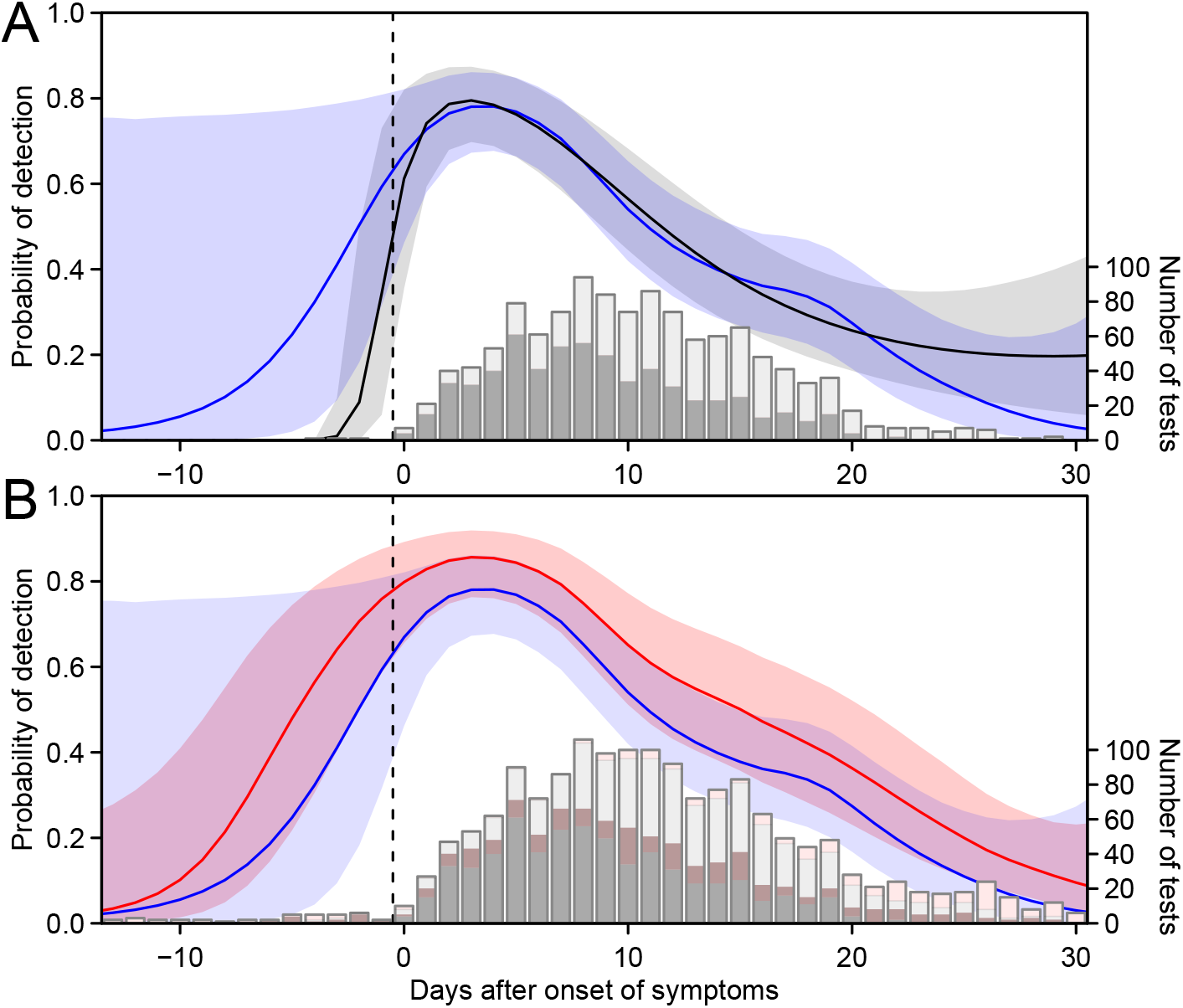
Longitudinal predictions of the detection rate of RT-qPCR testing for SARS-CoV-2. A) Comparison between polynomial and Bayesian autoregressive moving average state space models (B-ARMA-SSM) of SARS-COV-2 detection rates. Bars show positive (dark) and negative (light) test results from previously published studies of RT-qPCR collected by Kucirka et al. ^1^ Curves show the SARS-CoV-2 detection probabilities estimated by polynomial regression (black) and B-ARMA-SSM (blue) and their 95% credible intervals (shading). B) Comparison between the estimates of RT-qPCR detection rates before and after the inclusion of additional recently published data. Curves show detection probabilities estimated by B-ARMA-SSM before (blue, as in A) and after (red) the incorporation of data from seven recently published studies, including tests from 21 patients prior to the onset of symptoms. Shading shows 95% credible intervals. Bars show positive (dark) and negative (light) test results from the previous analysis and from the newly incorporated data (pink).

Fortunately, additional data has become available allowing a test of the predictive powers of these models. We identified 7 additional studies ^2,3,12–16^ by following the PubMed search strategy described previously ^1^. These new data contained 381 results from qPCR testing of 124 patients, including 21 patients measured prior to symptom onset. Combined with the previous seven studies, these data contained the results of 1619 RT-qPCR tests.

Fitting the B-ARMA-SSM to this combined data produced an updated estimate with remarkable similarity to its previous predictions (Figure 2). In addition, the estimates fell well within the credible intervals previously predicted by the model suggesting that the B-ARMA-SSM was able to adequately account for uncertainty in the data. In contrast, the new data did not seem to agree with the predictions made by the polynomial model. For example, the polynomial model predicted a 0% probability of detection earlier than 3 days prior to symptom onset yet 8 out of 13 tests administered between 4–7 days prior to symptom onset were positive (Figure 2).

Overall, the B-ARMA-SSM estimated that in the combined data RT-qPCR detection rates averaged 80% (95% CrI: 65–89%) on the day of symptom onset, increased to 86% (95% CrI: 76-92%) at three days after symptoms and then fell steadily further into infection. Similarly, detection was estimated as increasingly unlikely the earlier a patient was sampled prior to the onset of symptoms. For example, at 4 days prior to symptom onset, the average detection rate was estimated at 56% (95% CrI: 30-79%) and at 8 days prior to symptom onset, rates were estimated at 21% (95% CrI: 5-55%). Thus, detection of SARS-CoV-2 in presymptomatic individuals must be approached cautiously but is not nearly as difficult as previously suggested.

The relatively high false negative rates in SARS-CoV-2 RT-qPCR testing predicted here and previously ^1^ is worrisome. Of course, what is a “false negative” depends greatly on the purpose of testing. If tests are intended to provide documentation of previous SARS-CoV-2 infection then negative tests late in infection do indeed represent false negatives. However with the rapid development of serological testing, the more likely use for RT-qPCR testing is to detect positive cases prior to the onset of symptoms and to monitor viral clearance after symptom onset. In these cases, many “false negatives” are likely to be due to the reduction viral load to undetectable, and potentially less transmissible ^2,4,15,17,18^, levels rather than a failure of PCR testing. Thus at later time points, the false negative rate is likely overestimated. A similar problem arises in the interpretation of rates prior to symptom onset where it is unclear if a false negative is due to a missed test or instead because the patient has not yet been infected or reached significant levels of viral load.

In interpreting these data, it is also important to consider the limitations inherent to opportunistically collected data. The collected studies varied in techniques, assays and patient populations and some studies were estimated to have significantly higher or lower detection rates. For example, the van Kampen et al. ^15^ study was estimated to have the highest predicted detection rate with an estimated 97% (95% CrI: 94–95%) probability of detection in infected patients at three days after symptom onset. In addition, these analyses ignore censoring in the patient data. If a patient tests negative several times in a row then their doctor will often not continue to test them which would bias late stage data towards towards patients with prolonged disease. However, patient death or loss to followup due to more extreme disease progression could have an opposing effect. Further data from prospective sampling of at risk patients is essential to further characterize this critical period.

Population-wide screening efforts are being implemented in an attempt to reduce spread from asymptomatic and presymptomatic individuals infected with SARS-CoV-2. An understanding of detection rates is critical to these efforts. The preliminary analyses presented here suggest that, although testing will certainly miss some infections, the detection of many presymptomatic individuals is indeed possible and offers the potential to greatly reduce the spread of SARS-CoV-2^19**?**^.

## Methods

Data from seven studies ^5–11^ analyzed previously was obtained from Kucirka et al. ^1^. Additional data was collected from seven additional studies reporting testing in patients who developed symptoms, were tested multiple times and tested positive at least once ^2,3,12–16^. Where raw data were unavailable, data were digitized from published plots. Data from Arons et al. ^2^ was provided by personal communication. Only nasopharyngeal swabs were used from Xiao et al. ^12^ and tests detecting ≥ 1 PCR target were counted as positives. Only upper respiratory tract samples were used from van Kampen et al. ^15^ The data from a preprint by Kujawski et al. ^8^ in the initial data was replaced by data from the final publication by COVID-19 Investigation Team ^20^

In order to estimate the longitudinal progression of detection rates in SARS-CoV-2 patients, we developed a Bayesian autoregressive moving average state space model (B-ARMA-SSM). This model attempts to reduce potential misinterpretations in data sparse time periods by assuming simply that the detection rate on a given day should on average resemble that of the previous day and that increases or decreases in detection rate on a previous day will on average tend to continue. These probability are not directly observed but are inferred from the counts of positive and negative tests. To anchor the model, detection at 20 days prior to symptom onset is given a low prior probability. Differential detection rates in the individual studies are modeled as a constant multiplicative change in odds ratio drawn from a normal distribution of potential study offsets. The probabilities of RT-qPCR detection were thus:

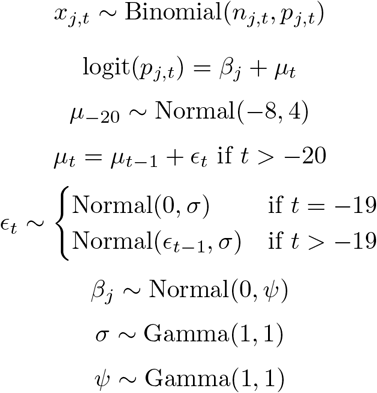

where *x*_*j,t*_ is the number of positives out of *n*_*j,t*_ total tests from study *j* at *t* days after symptom onset with estimated binomial probability *p*_*j,t*_ and study effect *B*_*j*_. The posterior probabilities of the Bayesian model were estimated using Markov chain Monte Carlo sampling as implemented in Stan ^21^ and analysis and plotting were performed in R ^22^.

## Data Availability

All data and code is available at https://github.com/sherrillmix/covidRTPCR and will be archived at Zenodo prior to final publication.

## References

[1] Lauren M. Kucirka, Stephen A. Lauer, Oliver Laeyendecker, Denali Boon, and Justin Lessler. Variation in false-negative rate of reverse transcriptase polymerase chain reaction-based SARS-CoV-2 tests by time since exposure. Ann Intern Med, May 2020. ISSN 1539-3704. doi: 10.7326/M20-1495.

[2] Melissa M. Arons, Kelly M. Hatfield, Sujan C. Reddy, Anne Kimball, Allison James, Jesica R. Jacobs, Joanne Taylor, Kevin Spicer, Ana C. Bardossy, Lisa P. Oakley, Sukarma Tanwar, Jonathan W. Dyal, Josh Harney, Zeshan Chisty, Jeneita M. Bell, Mark Methner, Prabasaj Paul, Christina M. Carlson, Heather P. McLaughlin, Natalie Thornburg, Suxiang Tong, Azaibi Tamin, Ying Tao, Anna Uehara, Jennifer Harcourt, Shauna Clark, Claire Brostrom-Smith, Libby C. Page, Meagan Kay, James Lewis, Patty Montgomery, Nimalie D. Stone, Thomas A. Clark, Margaret A. Honein, Jeffrey S. Duchin, and John A. Jernigan. Presymptomatic SARS-CoV-2 infections and transmission in a skilled nursing facility. N Engl J Med, April 2020. ISSN 1533-4406. doi: 10.1056/NEJMoa2008457.

[3] Seong Eun Kim, Hae Seong Jeong, Yohan Yu, Sung Un Shin, Soosung Kim, Tae Hoon Oh, Uh Jin Kim, Seung-Ji Kang, Hee-Chang Jang, Sook-In Jung, and Kyung-Hwa Park. Viral kinetics of SARS-CoV-2 in asymptomatic carriers and presymptomatic patients. Int J Infect Dis, 95:441–443, May 2020. ISSN 1878-3511. doi: 10.1016/j.ijid.2020.04.083.

[4] Xi He, Eric HY Lau, Peng Wu, Xilong Deng, Jian Wang, Xinxin Hao, Yiu Chung Lau, Jessica Y Wong, Yujuan Guan, Xinghua Tan, et al. Temporal dynamics in viral shedding and transmissibility of COVID-19. Nat Med, 26(5):672–675, 2020. doi: 10.1101/2020.03.15.20036707.

[5] Kostas Danis, Olivier Epaulard, Thomas Bénet, Alexandre Gaymard, Séphora Campoy, Elisabeth Bothelo-Nevers, Maude Bouscambert-Duchamp, Guillaume Spaccaferri, Florence Ader, Alexandra Mailles, Zoubida Boudalaa, Violaine Tolsma, Julien Berra, Sophie Vaux, Emmanuel Forestier, Caroline Landelle, Erica Fougere, Alexandra Thabuis, Philippe Berthelot, Raphael Veil, Daniel Levy-Bruhl, Christian Chidiac, Bruno Lina, Bruno Coignard, Christine Saura, and Investigation Team. Cluster of coronavirus disease 2019 (COVID-19) in the French Alps, 2020. Clin Infect Dis, April 2020. ISSN 1537-6591. doi: 10.1093/cid/ciaa424.

[6] Roman Wölfel, Victor M. Corman, Wolfgang Guggemos, Michael Seilmaier, Sabine Zange Marcel A. Müller, Daniela Niemeyer, Terry C. Jones, Patrick Vollmar, Camilla Rothe, Michael Hoelscher, Tobias Bleicker, Sebastian Brünink, Julia Schneider, Rosina Ehmann, Katrin Zwirglmaier, Christian Drosten, and Clemens Wendtner. Virological assessment of hospitalized patients with COVID-2019. Nature, 581: 465–469, May 2020. ISSN 1476-4687. doi: 10.1038/s41586-020-2196-x.

[7] Eu Suk Kim, Bum Sik Chin, Chang Kyung Kang, Nam Joong Kim, Yu Min Kang, Jae Phil Choi, Dong Hyun Oh, Jeong Han Kim, Boram Koh, Seong Eun Kim, Na Ra Yun, Jae Hoon Lee, Jin Yong Kim, Yeonjae Kim, Ji Hwan Bang, Kyoung Ho Song, Hong Bin Kim, Ki Hyun Chung, Myoung Don Oh, and Korea National Committee for Clinical Management of COVID-19. Clinical course and outcomes of patients with severe acute respiratory syndrome coronavirus 2 infection: a preliminary report of the first 28 patients from the Korean cohort study on COVID-19. J Korean Med Sci, 35:e142, April 2020. ISSN 1598-6357. doi: 10.3346/jkms.2020.35.e142.

[8] Stephanie A Kujawski, Karen K Wong, Jennifer P Collins, Lauren Epstein, Marie E Killerby, Claire M Midgley, Glen R Abedi, N Seema Ahmed, Olivia Almendares, Francisco N Alvarez, et al. First 12 patients with coronavirus disease 2019 (COVID-19) in the united states. MedRxiv, 2020.

[9] Juanjuan Zhao, Quan Yuan, Haiyan Wang, Wei Liu, Xuejiao Liao, Yingying Su, Xin Wang, Jing Yuan, Tingdong Li, Jinxiu Li, Shen Qian, Congming Hong, Fuxiang Wang, Yingxia Liu, Zhaoqin Wang, Qing He, Zhiyong Li, Bin He, Tianying Zhang, Yang Fu, Shengxiang Ge, Lei Liu, Jun Zhang, Ningshao Xia, and Zheng Zhang. Antibody responses to SARS-CoV-2 in patients of novel coronavirus disease 2019. Clin Infect Dis, March 2020. ISSN 1537-6591. doi: 10.1093/cid/ciaa344.

[10] Lei Liu, Wanbing Liu, Yaqiong Zheng, Xiaojing Jiang, Guomei Kou, Jinya Ding, Qiongshu Wang, Qianchuan Huang, Yinjuan Ding, Wenxu Ni, Wanlei Wu, Shi Tang, Li Tan, Zhenhong Hu, Weitian Xu, Yong Zhang, Bo Zhang, Zhongzhi Tang, Xinhua Zhang, Honghua Li, Zhiguo Rao, Hui Jiang, Xingfeng Ren, Shengdian Wang, and Shangen Zheng. A preliminary study on serological assay for severe acute respiratory syndrome coronavirus 2 (SARS-CoV-2) in 238 admitted hospital patients. Microbes Infect, 22:206–211, 2020. ISSN 1769-714X. doi: 10.1016/j.micinf.2020.05.008.

[11] Li Guo, Lili Ren, Siyuan Yang, Meng Xiao, D. Chang, Fan Yang, Charles S. Dela Cruz, Yingying Wang, Chao Wu, Yan Xiao, Lulu Zhang, Lianlian Han, Shengyuan Dang, Yan Xu, Qiwen Yang, Shengyong Xu, Huadong Zhu, Yingchun Xu, Qi Jin, Lokesh Sharma, Linghang Wang, and Jianwei Wang. Profiling early humoral response to diagnose novel coronavirus disease (COVID-19). Clin Infect Dis, March 2020. ISSN 1537-6591. doi: 10.1093/cid/ciaa310.

[12] Ai Tang Xiao, Yi Xin Tong, and Sheng Zhang. Profile of RT-PCR for SARS-CoV-2: a preliminary study from 56 COVID-19 patients. Clin Infect Dis, April 2020. ISSN 1537-6591. doi: 10.1093/cid/ciaa460.

[13] Jonathan Altamirano, Prasanthi Govindarajan, Andra L. Blomkalns, Lauren E. Kushner, Bryan Andrew Stevens, Benjamin A. Pinsky, and Yvonne Maldonado. Assessment of sensitivity and specificity of patient-collected lower nasal specimens for sudden acute respiratory syndrome coronavirus 2 testing. JAMA Netw Open, 3:e2012005, June 2020. ISSN 2574-3805. doi: 10.1001/jamanetworkopen.2020.12005.

[14] Ivan Fan-Ngai Hung, Vincent Chi-Chung Cheng, Xin Li, Anthony Raymond Tam, Derek Ling-Lung Hung, Kelvin Hei-Yeung Chiu, Cyril Chik-Yan Yip, Jian-Piao Cai, Deborah Tip-Yin Ho, Shuk-Ching Wong, Sally Sau-Man Leung, Man-Yee Chu, Milky Oi-Yan Tang, Jonathan Hon-Kwan Chen, Rosana Wing-Shan Poon, Agnes Yim-Fong Fung, Ricky Ruiqi Zhang, Erica Yuen-Wing Yan, Lin-Lei Chen, Charlotte Yee-Ki Choi, Kit-Hang Leung, Tom Wai-Hin Chung, Sonia Hiu-Yin Lam, Tina Poy-Wing Lam, Jasper Fuk-Woo Chan, Kwok-Hung Chan, Tak-Chiu Wu, Pak-Leung Ho, Johnny Wai-Man Chan, Chak-Sing Lau, Kelvin Kai-Wang To, and Kwok-Yung Yuen. SARS-CoV-2 shedding and seroconversion among passengers quarantined after disembarking a cruise ship: a case series. Lancet Infect Dis, June 2020. ISSN 1474-4457. doi: 10.1016/S1473-3099(20)30364-9.

[15] Jeroen JA van Kampen, David AMC van de Vijver, Pieter LA Fraaij, Bart L Haagmans, Mart M Lamers, Nisreen Okba, Johannes PC van den Akker, Henrik Endeman, Diederik AMPJ Gommers, Jan J Cornelissen, et al. Shedding of infectious virus in hospitalized patients with coronavirus disease-2019 (COVID-19): duration and key determinants. medRxiv, 2020. doi: 10.1101/2020.06.08.20125310.

[16] Wang-Da Liu, Sui-Yuan Chang, Jann-Tay Wang, Ming-Jui Tsai, Chien-Ching Hung, Chia-Lin Hsu, and Shan-Chwen Chang. Prolonged virus shedding even after seroconversion in a patient with COVID-19. J Infect, 2020. doi: 10.1016/j.jinf.2020.03.063.

[17] Jared Bullard, Kerry Dust, Duane Funk, James E. Strong, David Alexander, Lauren Garnett, Carl Boodman, Alexander Bello, Adam Hedley, Zachary Schiffman, Kaylie Doan, Nathalie Bastien, Yan Li, Paul G. Van Caeseele, and Guillaume Poliquin. Predicting infectious SARS-CoV-2 from diagnostic samples. Clin Infect Dis, May 2020. ISSN 1537-6591. doi: 10.1093/cid/ciaa638.

[18] Sin Fun Sia, Li-Meng Yan, Alex WH Chin, Kevin Fung, Ka-Tim Choy, Alvina YL Wong, Prathanporn Kaewpreedee, Ranawaka APM Perera, Leo LM Poon, John M Nicholls, et al. Pathogenesis and transmission of SARS-CoV-2 in golden hamsters. Nature, pages 1–7, 2020. doi: 10.1038/s41586-020-2342-5.

[19] Seyed M. Moghadas, Meagan C. Fitzpatrick, Pratha Sah, Abhishek Pandey, Affan Shoukat, Burton H. Singer, and Alison P. Galvani. The implications of silent transmission for the control of COVID-19 outbreaks. Proc Natl Acad Sci U S A, July 2020. ISSN 1091-6490. doi: 10.1073/pnas.2008373117.

[20] COVID-19 Investigation Team. Clinical and virologic characteristics of the first 12 patients with coronavirus disease 2019 (COVID-19) in the United States. Nat Med, April 2020. ISSN 1546-170X. doi: 10.1038/s41591-020-0877-5.

[21] Bob Carpenter, Andrew Gelman, Matthew D Hoffman, Daniel Lee, Ben Goodrich, Michael Betancourt, Marcus Brubaker, Jiqiang Guo, Peter Li, and Allen Riddell. Stan: A probabilistic programming language. J of Stat Softw, 76(1), 2017. doi: 10.18637/jss.v076.i01.

[22] R Core Team. R: A Language and Environment for Statistical Computing. R Foundation for Statistical Computing, Vienna, Austria, 2018. URL https://www.R-project.org/.

